# Overprotection and overcontrol in childhood: An evaluation on reliability and validity of 33-item Expanded Childhood Trauma Questionnaire (CTQ-33), Chinese version

**DOI:** 10.1101/2021.11.15.21266378

**Authors:** Zhipeng Wu, Zhening Liu, Zhengqian Jiang, Xingzi Fu, Qian Deng, Lena Palaniyappan, Zhibiao Xiang, Danqing Huang, Yicheng Long

**Author notes:** **Correspondence:** Yicheng Long.

## Abstract

Overprotection and overcontrol from parents or other family members, which are not rare in the Chinese culture, have been suggested to be traumatic experiences for some children. However, research on overprotection/overcontrol is much rarer in China compared with other childhood trauma subtypes. One of the possible reasons for this is the lack of easy and feasible screening tools. In this study, we therefore translated and validated a Chinese version of the 33-item Childhood Trauma Questionnaire (CTQ-33), which was expanded from the widely-used 28-item CTQ with an additional overprotection/overcontrol subscale. A total of 248 young healthy participants were recruited and completed the Chinese version of CTQ-33, and 50 of them were retested after an interval of two weeks. At baseline, all participants also completed the 9-item Patient Health Questionnaire and the 7-item Generalized Anxiety Disorder Scale to assess their depression and anxiety, respectively. Our main findings include that: (1) the Chinese version of CTQ-33 showed a good internal consistency (Cronbach’s α coefficient = 0.733) and an excellent test-retest reliability over a two-week period (ICC = 0.861); (2) the previously reported significant associations between the overprotection/overcontrol and other subtypes of childhood trauma (abuse and neglect), as well as psychopathological conditions such as depression can all be replicated using the Chinese version of CTQ-33. These results suggest that the Chinese version of CTQ-33 would be a promising tool for assessing various subtypes of childhood adversities, especially the overprotection/overcontrol experiences in Chinese populations.

## 1 Introduction

Over the past two decades, research on the impact of childhood trauma on mental health has notably expanded (Carr et al., 2013; Maschi et al., 2013). There is now ample evidence demonstrating an association between childhood trauma and psychotic-like experiences (Sun et al., 2017b) as well as adult-onset psychiatric disorders such as schizophrenia (Carr et al., 2013; Wang et al., 2013), bipolar disorder (Watson et al., 2014) and major depressive disorder (Wiersma et al., 2009). Given its role in the development of common psychiatric disorders, studying childhood trauma is seen as critical for the integrative “bio-psycho-social” model (Read et al., 2005; Sideli et al., 2020).

Childhood trauma is not a single entity; different trauma subtypes may lead to differential long-term effects on the human brain (Cassiers et al., 2018; Li et al., 2021). One of the most well studied approach to distinguish subtypes of childhood trauma events is categorizing it into five factors: emotional abuse, physical abuse, sexual abuse, physical neglect, and emotional neglect (Carr et al., 2013; Cassiers et al., 2018; Şar et al., 2004). These are often assessed by the 28-item Childhood Trauma Questionnaire (CTQ-28) (Bernstein et al., 2003; Huang et al., 2021; Tetik et al., 2020; Xiang et al., 2021). However, some recent observations indicate that other important negative childhood events are equally traumatic but cannot be classified into any of the above five factors (Şar, 2020). One example is overprotection/overcontrol from parents or other family members, which was proved to be a risk factor for multiple mental problems/diseases (Azar et al., 2007; Herbert and Dahlquist, 2008; Yoshida et al., 2005) and has attracted attentions in recent psychological studies (Farina et al., 2021; Jones et al., 2021; Şar et al., 2021; Wu et al., 2020). Thus, to provide a promising tool for better differentiation of various types of childhood trauma experiences, Şar et al. (Şar et al., 2021) developed an expanded 33-item Childhood Trauma Questionnaire (CTQ-33) on the basis of conventional CTQ-28, with overprotection/overcontrol as an additional factor. The CTQ-33 has been proved to be reliable and valid in both non-clinical participants and patients with psychiatric disorders, with both English and Turkish versions provided (Şar et al., 2021).

Similar to countries in other cultural contexts, perceived parenting styles with overprotection and overcontrol are not rare in China (Mousavi et al., 2016). To our knowledge, however, research on overprotection/overcontrol is much rarer compared with other childhood trauma subtypes (abuse and neglect) in China. One of the potential reasons is the lacking of a publicly available, feasible and easy assessment tool for childhood experiences of overprotection/overcontrol in Chinese language. A possible choice is the Short-form Egna Minnen av Barndoms Uppfostran (s-EMBU) (Arrindell et al., 1999), which includes items measuring “overprotection” and has been translated into Chinese (Li et al., 2012). However, the s-EMBU was designed to assess memories of parental rearing behaviors from all aspects (de Haas et al., 1994) rather than traumatic events, and doesn’t have a specific subscale for overcontrols. Another important limitation of s-EMBU and other similar assessments is that they only assess behaviors of one’s parents but ignore other family members, while there are a large number of “left-behind children” in China who are cared for by relatives instead of their parents (Sun et al., 2017a). Thus, developing and validating the Chinese version of CTQ-33 might help filling these gaps and make contributions to a more comprehensive assessment of childhood trauma in Chinese culture.

For the above reasons, this study aimed to translate and validate a Chinese version of CTQ-33. In specific, after translating, we evaluated the reliability and validity of CTQ-33 by (1) measuring its internal consistency and test-retest reliability in a group of Chinese young adults; and (2) seeing if the previously reported significant correlations between overprotection/overcontrol and other subtypes of childhood trauma in CTQ-33, as well as other psychopathological conditions (e.g., depression) (Şar et al., 2021) would be replicated in Chinese participants. We hypothesized that the Chinese version of CTQ-33 would have good internal consistency and test-retest reliability, and the significant associations between overprotection/overcontrol and other childhood trauma subtypes as well as other psychopathological conditions would be replicated in Chinese populations.

## 2 Methods

### 2.1 Participants

The participants included in this study were 248 young healthy adults who were either undergraduate or postgraduate students recruited from the Xiangya School of Medicine, Changsha, Hunan, China. All participants had no history of any psychiatric disorder or other severe illness. Among the participants, there were 88 males and 160 females; the average age was 20.56 ± 2.22 (mean ± standard deviation) years. From July 19^th^, 2021 to July 22^nd^, 2021, all participants completed the baseline assessments of CTQ-33 and other scales using the largest online survey platform in China, “Questionnaire Star” (www.wjx.cn). From these participants, 50 subjects were randomly selected and asked to re-complete the CTQ-33 after an interval of two weeks. There were no significant differences in age, sex and scores of most assessments between the participants who were retested and those who were not (**Table 1**). The study was approved by the Ethics Committees of the Second Xiangya Hospital of Central South University, Changsha, and all participants signed informed consents.

**Table 1.** Demographic and clinical characteristics of the participants at baseline. Between-group comparison were performed by t-test (for age), Chi-square test (for sex) or Mann-Whitney U test (for all clinical scales).

|  | All<br>participants (*n*<br>= 248) | Participants<br>who were<br>retested (*n* =<br>50) | Participants<br>who were<br>not retested<br>(*n* = 198) | Comparisons<br>between<br>participants<br>who<br>were/were<br>not retested |
| --- | --- | --- | --- | --- |
| Sex (male/female) | 88/160 | 21/29 | 67/131 | $\chi^2 = 1.162, p = 0.281$ |
| Age (years) | 20.56 ± 2.22 | 20.46 ± 1.31 | 20.59 ± 2.40 | $t = 0.372, p = 0.710$ |
| PHQ-9 score | 5.88 ± 4.40 | 5.70 ± 3.55 | 5.92 ± 4.60 | $p = 0.869$ |
| GAD-7 score | 4.20 ± 3.71 | 4.18 ± 3.05 | 4.21 ± 3.87 | $p = 0.614$ |
| CTQ-33 total score | 45.65 ± 9.95 | 45.66 ± 9.78 | 45.65 ±<br>10.02 | $p = 0.969$ |
| CTQ-33 subscale<br>scores |  |  |  |  |
| Physical abuse | 5.58 ± 1.30 | 5.72 ± 1.80 | 5.55 ± 1.15 | $p = 0.820$ |
| Emotional neglect | 9.64 ± 3.76 | 9.88 ± 3.89 | 9.58 ± 3.73 | $p = 0.690$ |
| Emotional abuse | 7.06 ± 2.33 | 7.44 ± 2.19 | 6.96 ± 2.36 | $p = 0.032$ |
| Physical neglect | 7.84 ± 2.65 | 7.90 ± 2.39 | 7.82 ± 2.71 | $p = 0.554$ |
| Sexual abuse | 5.48 ± 1.42 | 5.40 ± 1.23 | 5.50 ± 1.47 | $p = 0.244$ |
| Overprotection/overcontrol | 10.05 ± 3.50 | 9.32 ± 2.67 | 10.23 ± 3.66 | $p = 0.206$ |
| Minimization/denial | 0.77 ± 0.92 | 0.70 ± 0.86 | 0.78 ± 0.93 | $p = 0.686$ |

### 2.2 Study design and assessments

All participants (*n* = 248) completed the translated Chinese version of CTQ-33 to assess childhood trauma, the 9-item Patient Health Questionnaire (PHQ-9) to assess depression and the 7-item Generalized Anxiety Disorder Scale (GAD-7) to assess anxiety at baseline. All internal consistency and correlation analyses were performed based on the data at baseline. Furthermore, 50 participants re-completed the CTQ-33 after two weeks of the baseline assessment to evaluate its test-retest reliability.

#### Chinese version of CTQ-33

The CTQ-33 was developed by Şar et al. (Şar et al., 2021) on the basis of CTQ-28, with an additional subscale of overprotection/overcontrol. The overprotection/overcontrol subscale includes five new items as below: “People in my family restricted my contacts with my peers and friend (item 29)”; “People in my family intervened with my personal matters (item 30)”; “My mother and father let me carry on tasks by my own (item 31, reversed item)”; “People in my family followed my life so closely that I felt intruded (item 32)”; and “My mother or father used to check me by digging through my personal belongings (item 33)”. Same as the other original items, all these five items are 5-point Likert-type questions. Respondents are required to rate each item from 1 to 5 according to the frequency (1 = never, 5 = very often for regular items; 1 = very often, 5 = never for reversed items). We translated these five new items into Chinese and added them to the previous Chinese version of CTQ-28 (Zhao et al., 2005) to form the Chinese version of CTQ-33. Translations were performed by the corresponding author of this article, Dr. Yicheng Long from Central South University. Chinese versions of the five new items were further translated back into English independently by a co-author, Ms. Danqing Huang. The original and back-translated items were then compared by another clinician, Mr. Zhibiao Xiang, to verify the consistency between them. The final translated Chinese versions of the five overprotection/overcontrol items can be found in **Supplemental Table 1**.

As a result, the CTQ-33 has six subscales: emotional abuse, physical abuse, sexual abuse, physical neglect emotional neglect and overprotection/overcontrol. Scores of each subscale were calculated by summing its five items and ranged from 5 to 25. Note that there are three minimization/denial items in the original CTQ-28 and they were retained in the CTQ-33. The minimization/denial score was calculated referring to previous literature (MacDonald et al., 2016). Translation and utilization of the CTQ-33 was approved by the original author, Prof. Vedat Şar (Şar et al., 2021) from the Koç University School of Medicine.

#### PHQ-9

the PHQ-9 is a 7-item self-administered screening instrument to assess depression during the past two weeks, with each item ranging from 0 to 3 (Kroenke et al., 2001; Spitzer et al., 1999). Chinese version of the PHQ-9 has been validated previously (Wang et al., 2014).

#### GAD-7

the GAD-7 is a 7-item self-report questionnaire to assess anxiety during the past two weeks, with each item ranging from 0 to 3 (Spitzer et al., 2006). Chinese version of the GAD-7 has been validated previously (Tong et al., 2016; Zeng et al., 2013; Zhang et al., 2021).

### 2.3 Internal consistency and test-retest reliability

Internal consistency of the CTQ-33 total/subscale scores were estimated by Cronbach’s α coefficients, with a Cronbach’s α coefficient > 0.7 indicating good internal consistency (McHorney and Tarlov, 1995; Wu et al., 2021a; Zhang et al., 2021). Internal consistency of the PHQ-9 and GAD-7 was also tested by the same method. Two-week test-retest reliability of the CTQ-33 total/subscale scores were estimated by two-way random effects intra-class correlation coefficients (ICCs) (Shrout and Fleiss, 1979). According to ICCs, the test-retest reliabilities can be interpreted as poor (ICC < 0.4), moderate (0.4 ≤ ICC < 0.6), good (0.6 ≤ ICC < 0.75) and excellent (ICC ≥ 0.75) (Cao et al., 2019; Cicchetti, 1994; Long et al., 2021).

### 2.4 Correlations

Associations between the overprotection/overcontrol scores and scores of other subscales in the CTQ-33, as well as between the CTQ-33 total/subscale scores and PHQ-9/GAD-7 scores were estimated by Spearman rank correlations. Threshold for significance was set at *p* < 0.05.

### 2.5 Exploratory analysis on demographic-related differences

Since previous studies have widely reported the demographic-related differences, especially the sex-related differences in childhood trauma and other mental health problems (Eyuboglu et al., 2021; Holik et al., 2021; Lee and Feng, 2021; Thorisdottir et al., 2021; Wu et al., 2021b), we further performed an exploratory analysis on possible age- and sex-related differences in all tested scales. Here, they were (1) correlated with age by Spearman rank correlations; and (2) compared between the male and female participants by Mann-Whitney U tests. Statistical significances were set at *p* < 0.05.

## 3 Results

### 3.1 Internal consistency and test-retest reliability

The CTQ-33 showed a good internal consistency (Cronbach’s α coefficient = 0.733) and an excellent test-retest reliability (ICC = 0.861). Moreover, as shown in **Table 2**, most subscales of the CTQ-33 (including the new added overprotection/overcontrol subscale) had good internal consistencies (Cronbach’s α coefficient > 0.7) and all subscales had good (0.6 ≤ ICC < 0.75) or excellent (ICC ≥ 0.75) test-retest reliabilities. The PHQ-9 and GAD-7 also had good internal consistencies (Cronbach’s α coefficient = 0.867/0.894 for the PHQ-9/GAD-7).

**Table 2.** Internal consistency (measured by Cronbach’s α coefficients) and two-week test-retest reliability (measured by ICCs) of the CTQ-33 and its subscales.

| CTQ-33 subscale | Cronbach's $\alpha$<br>coefficient | ICC (with 95%<br>confidence intervals) |
| --- | --- | --- |
| Physical abuse | 0.739 | 0.869 (0.781, 0.924) |
| Emotional neglect | 0.780 | 0.814 (0.694, 0.890) |
| Emotional abuse | 0.627 | 0.841 (0.737, 0.907) |
| Physical neglect | 0.459 | 0.616 (0.403, 0.764) |
| Sexual abuse | 0.855 | 0.876 (0.793, 0.928) |
| Overprotection/overcontrol | 0.804 | 0.720 (0.554, 0.831) |
| Minimization/<br>denial | 0.478 | 0.610 (0.402, 0.759) |
| Total | 0.733 | 0.861 (0.786, 0.919) |

### 3.2 Correlations

Significant positive correlations were found between most pairs of subscales in the CTQ-33 (*p* < 0.05, **Table 3**). Moreover, significant positive correlations were found between the PHQ-9/GAD-7 scores and CTQ-33 total score, as well as between the PHQ-9/GAD-7 scores and scores of all subscales in the CTQ-33 (*p* < 0.05, **Table 4**).

**Table 3.**
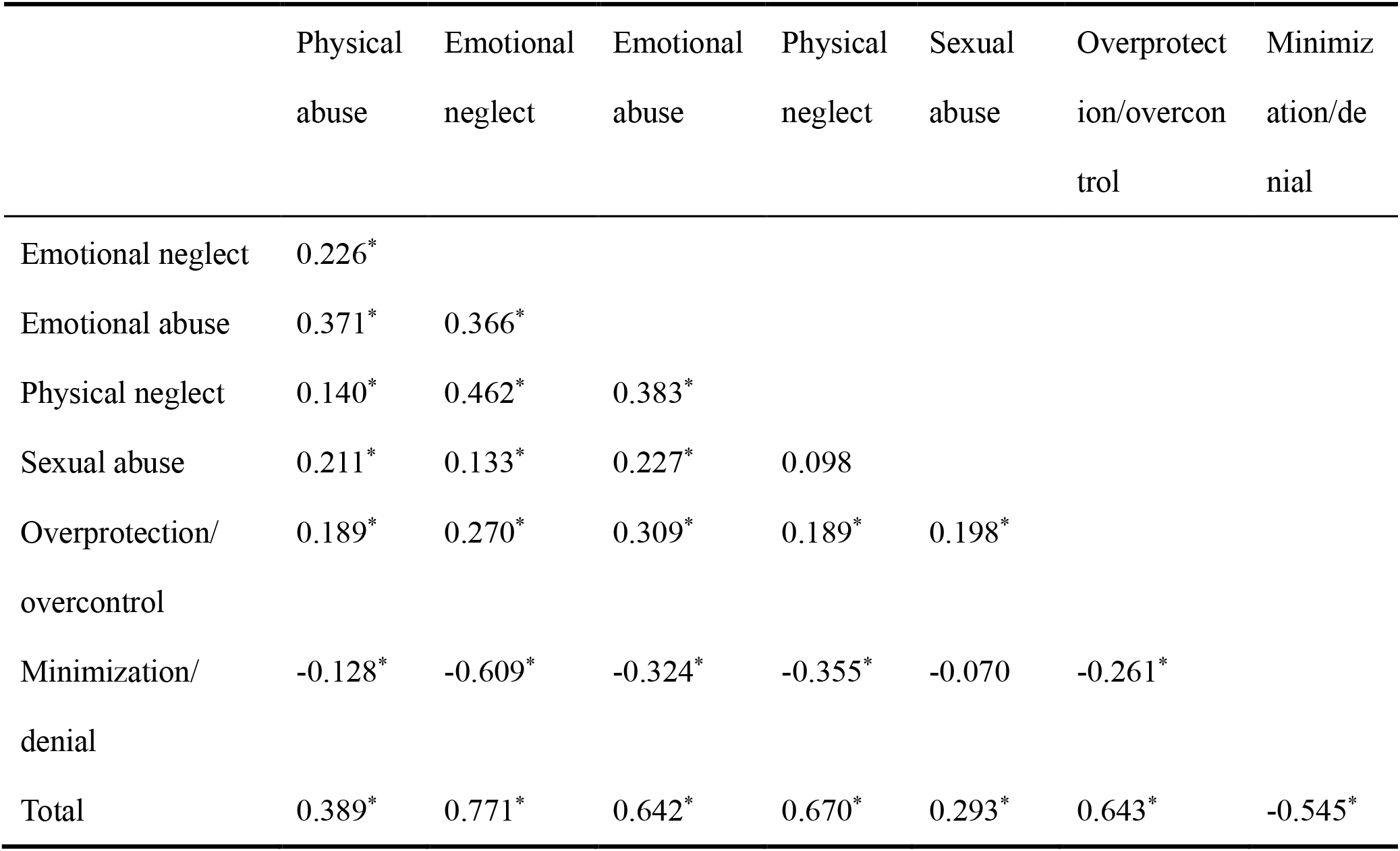
Spearman correlation coefficients between subscale scores of the CTQ-33. The “*” indicates a significant correlation (*p* < 0.05).

|  | Physical<br>abuse | Emotional<br>neglect | Emotional<br>abuse | Physical<br>neglect | Sexual<br>abuse | Overprotect<br>ion/overcon<br>trol | Minimiz<br>ation/de<br>nial |
| --- | --- | --- | --- | --- | --- | --- | --- |
| Emotional neglect | 0.226* |  |  |  |  |  |  |
| Emotional abuse | 0.371* | 0.366* |  |  |  |  |  |
| Physical neglect | 0.140* | 0.462* | 0.383* |  |  |  |  |
| Sexual abuse | 0.211* | 0.133* | 0.227* | 0.098 |  |  |  |
| Overprotection/<br>overcontrol | 0.189* | 0.270* | 0.309* | 0.189* | 0.198* |  |  |
| Minimization/<br>denial | -0.128* | -0.609* | -0.324* | -0.355* | -0.070 | -0.261* |  |
| Total | 0.389* | 0.771* | 0.642* | 0.670* | 0.293* | 0.643* | -0.545* |

**Table 4.** Spearman correlation coefficients between the CTQ-33 subscale scores and PHQ-9/GAD-7 scores. The “*” indicates a significant correlation (*p* < 0.05).

|  | PHQ-9 | GAD-7 |
| --- | --- | --- |
| Physical abuse | 0.241* | 0.131* |
| Emotional neglect | 0.353* | 0.236* |
| Emotional abuse | 0.435* | 0.288* |
| Physical neglect | 0.264* | 0.190* |
| Sexual abuse | 0.251* | 0.129* |
| Overprotection/<br>overcontrol | 0.376* | 0.315* |
| Minimization/<br>denial | -0.225* | -0.175* |
| Total | 0.515* | 0.352* |

### 3.3 Demographic-related differences

There were no significant correlations between age and scores of all adopted scales or subscales (all *p* > 0.05). The males had a significantly higher physical abuse score than females (*p* < 0.001, **Figure 1**), while no significant sex differences were found in other scales/subscales (all *p* > 0.05).

**Figure 1.**
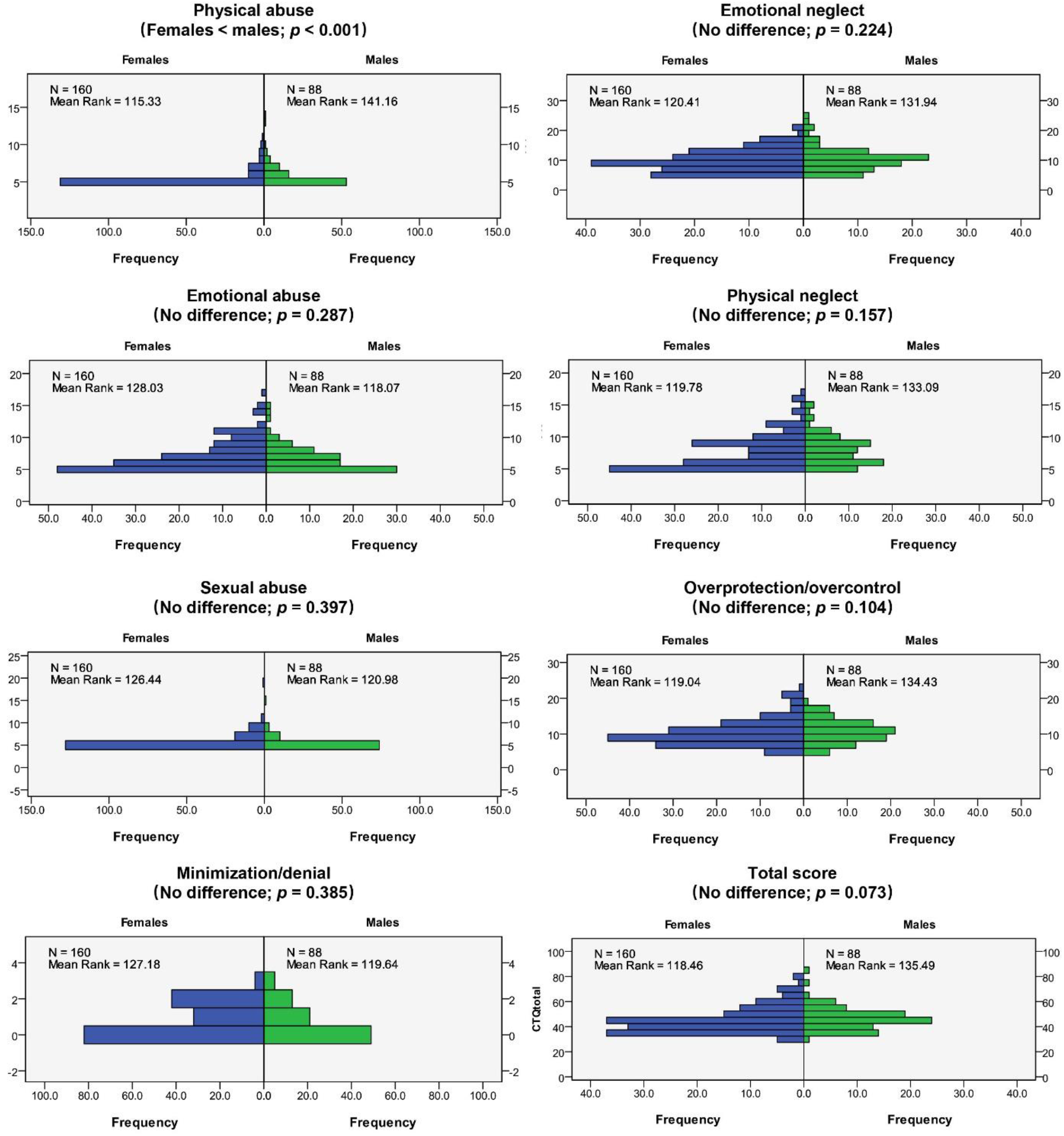
Comparisons on the CTQ-33 total/subscale scores between the female and male participants.

## 4 Discussion

In this study, we translated and validated a Chinese version of CTQ-33, which was expanded from the original CTQ-28 with an additional subscale of overprotection/overcontrol. Our main results include: (1) the Chinese version of CTQ-33 showed a good internal consistency and an excellent test-retest reliability over two weeks; (2) the previously reported significant associations between the overprotection/overcontrol and other subtypes of childhood trauma (abuse and neglect), as well as common psychopathological conditions including depression/anxiety can all be replicated using the Chinese version of CTQ-33.

The first main conclusion suggested by this work is that the Chinese version of CTQ-33 is a reliable instrument with a good internal consistency (Cronbach’s α coefficient = 0.733) and an excellent test-retest reliability (ICC = 0.861 when retested over two weeks). The CTQ-33 was expanded from the conventional CTQ-28 by Şar et al. (Şar et al., 2021) with an additional factor of overprotection/overcontrol behaviors from parents or other family members. To the best of our knowledge, while the CTQ-33 has been validated with both English and Turkish versions provided (Şar et al., 2021), this is the first study to examine the reliability and validity of its Chinese version. Overprotection/overcontrol rearing behaviors from parents and other family members are not rare in China (Mousavi et al., 2016), and they were suggested to be traumatic childhood experiences (Şar, 2020). However, such a type of events was generally not included in the widely-used CTQ-28, or other assessments of childhood trauma histories which a Chinese version (J. J. Chang et al., 2021; Huang et al., 2021; Li et al., 2021; Xiang et al., 2021). Here, the Chinese version of CTQ-33 may fill such gaps, providing a promising assessment of overprotection/overcontrol behaviors in future studies on childhood adversity.

All subscales of the Chinese version of CTQ-33 had good internal consistencies (Cronbach’s α coefficient > 0.7), with good test-retest reliabilities (ICC > 0.6) found for most subscales (**Table 2**). The exceptions included the physical neglect subscale (Cronbach’s α coefficient = 0.459) and minimization/denial items (Cronbach’s α coefficient = 0.478), which was included in the original Chinese version of CTQ-28 (Zhao et al., 2005) and preserved in the new CTQ-33. The result of a relatively low internal consistency of the physical neglect subscale was consistent with multiple previous studies on the Chinese version of CTQ-28 (Cheng et al., 2018; Jiang et al., 2018). One possible reason is the differences in the definitions of physical neglect across different cultures; for example, in general Chinese parents agree with the adage “spare the rod and spoil the child”; but parents from other countries may not share the same view (Jiang et al., 2018). Besides the physical neglect subscale, the three minimization/denial items also showed a relatively low internal consistency, which is in line with previous research (Xiang et al., 2021). Therefore, the same as in the CTQ-28, it should be more cautious to use the physical neglect and minimization/denial scores compared with other subscales in the Chinese version of CTQ-33. Nevertheless, good internal consistencies and test-retest reliabilities were shown for the CTQ-33 as a whole and specially, for the additional overprotection/overcontrol subscale in the CTQ-33, suggesting that the new added items are as reliable as the previous version.

Highly consistent with the previous report by Şar et al. (Şar et al., 2021), significant positive relationships were found between the overprotection/overcontrol score and scores of all other childhood trauma subtypes in the CTQ-33 (**Table 3**) in the current study. Such results may further reinforce the opinion that overprotection/overcontrol is a kind of childhood traumatic experience, and also indicate that the CTQ-33 did not extensively deviate from the original version of CTQ-28 with an additional overprotection/overcontrol factor (Şar et al., 2021). Furthermore, the previously reported significant associations between participants’ overprotection/overcontrol scores and common psychopathological conditions such as depression (Şar et al., 2021) were also replicated in this study (**Table 4**). This replication, in part, validates the Chinese version of CTQ-33. Furthermore, among the various subscales that related to anxiety scores, the largest effect size related to overprotection/overcontrol, highlighting the importance of measuring this construct independently. Taken together, CTQ-33 Chinese version can provide critical information on childhood antecedents of mental illness during adulthood (Abraham et al., 2021; C. C. Chang et al., 2021).

In this study, we also explored the possible sex- and age-related effects on CTQ-33 scores. It was found that the CTQ-33 total score and scores of most of its subscales were not significantly related to age/sex, except a higher physical abuse score in males (**Figure 1**) which has been reported in previous studies (Fang et al., 2015; Lee and Feng, 2021). However, it should be noted that participants in this study were all young adults, and it may be needed to confirm such effects in populations with a broader range of age.

There are a number of limitations and future directions to be considered. First, the sample size in this study is relatively small. Further studies in a bigger sample to investigate the prevalence of overprotection/overcontrol experiences in Chinese population are needed and have been scheduled by our team. Second, this study was only performed in healthy participants. Future studies in clinical populations with mental disorders may provide more valuable information about the possible roles of overprotection/overcontrol in development of those diseases. Third, combing the CTQ-33 and biomedical technologies such as structural (Long et al., 2018) and functional (Long et al., 2020) neuroimaging methods in future studies may be a promising direction to expand our understanding of neural substrate underlying the long-terms effects of childhood overprotection/overcontrol experiences on one’s mental health.

## 5 Conclusions

In conclusion, this study translated and validated a Chinese version of CTQ-33, which was developed by expanding the widely-used CTQ-28 with an additional overprotection/overcontrol subscale. Our results showed that the Chinese version of CTQ-33 is reliable and valid with a good internal consistency and an excellent test-retest reliability. We also replicated the previously reported associations between the overprotection/overcontrol and other subtypes of childhood trauma, as well as depression/anxiety using the CTQ-33 in a Chinese population. These results suggest that the Chinese version of CTQ-33 would be a promising tool for assessing various subtypes of childhood adversities, especially the overprotection/overcontrol experiences in future research.

## Data Availability

All data produced in the present study are available upon reasonable request to the authors.

## Acknowledgments

We thank Prof. Vedat Şar from the Koç University School of Medicine for his approval to translate and utilize the CTQ-33 in our studies. We also would like to thank all students who served as research participants.

## Supplementary data

Supplementary material related to this article can be found in the online version.

### Appendix

**Supplemental Table 1.** The five additional items in the Chinese version of CTQ-33.

| Item | Original | Chinese translation |
| --- | --- | --- |
| 29 | People in my family restricted my contacts with my peers and friends. | 我的家人会限制我和朋友的交往。 |
| 30 | People in my family intervened with my personal matters. | 我的家人会干涉我的私事。 |
| 31 | My mother and father let me carry on tasks by my own (reversed item). | 我的父母会让我独立完成自己的事情。 |
| 32 | People in my family followed my life so closely that I felt intruded. | 我的家里人对我生活的关照有些过头了，让我觉得不舒服。 |
| 33 | My mother or father used to check me by digging through my personal belongings. | 我的父母经常随意查看我的私人物品。 |

